# Sample Pooling is a Viable Strategy for SARS-CoV-2 Detection in Low-Prevalence Settings

**DOI:** 10.1101/2020.08.26.20181719

**Authors:** Brian SW Chong, Thomas Tran, Julian Druce, Susan A Ballard, Julie A Simpson, Mike Catton

**Affiliations:** Victorian Infectious Diseases Reference Laboratory The Peter Doherty Institute for Infection and Immunity 792 Elizabeth St Melbourne VIC 3000 Australia; Microbiological Diagnostic Unit Public Health Laboratory The University of Melbourne at the Peter Doherty Institute for Infection and Immunity 792 Elizabeth St Melbourne VIC 3000 Australia; Centre for Epidemiology and Biostatistics Melbourne School of Population and Global Health The University of Melbourne Melbourne, Australia

## Abstract

**BACKGROUND:** The severe acute respiratory syndrome coronavirus 2 (SARS-CoV-2) pandemic has significantly increased demand on laboratory throughput and reagents for nucleic acid extraction and polymerase chain reaction (PCR). Reagent shortages may limit the expansion of testing required to scale back isolation measures.

**AIM:** To investigate the viability of sample pooling as a strategy for increasing test throughput and conserving PCR reagents; to report our early experience with pooling of clinical samples.

**METHODS:** A pre-implementation study was performed to assess the sensitivity and theoretical efficiency of two, four, and eight-sample pools in a real-time reverse transcription PCR-based workflow. A standard operating procedure was developed and implemented in two laboratories during periods of peak demand, inclusive of over 29,000 clinical samples processed in our laboratory.

**RESULTS:** Sensitivity decreased (mean absolute increase in cycle threshold value of 0.6, 2.3, and 3.0 for pools of two, four, and eight samples respectively) and efficiency increased as pool size increased. Gains from pooling diminished at high disease prevalence. Our standard operating procedure was successfully implemented across two laboratories. Increased workflow complexity imparts a higher risk of errors, and requires risk mitigation strategies. Turnaround time for individual samples increased, hence urgent samples should not be pooled.

**CONCLUSIONS:** Pooling is a viable strategy for high-throughput testing of SARS-CoV-2 in low-prevalence settings.

## 1. INTRODUCTION

Timely, scalable, and accurate diagnostic testing for SARS-CoV-2 underpins the public health response to the COVID-19 pandemic and clinical care of suspected cases. Real-time reverse transcription-polymerase chain reaction (rRT-PCR) on a respiratory sample – most commonly an upper respiratory tract swab – is the main diagnostic modality for detection of SARS-CoV-2 [1]. However, global shortages of PCR reagents and consumables have constrained testing capacity, and may continue to limit the expansion of testing that countries will need to safely scale back isolation measures. Group testing, or pooling of patient samples has previously been employed in mass screening, both for nucleic acid testing and immunoassays [2, 3]. This approach may increase the throughput of PCR and improve the utilisation of PCR reagents during difficult times for routine RT-PCR-based diagnostic workflow. A recent Californian study has described the process of pooling for SARS-CoV-2 testing on a relatively small number of samples, with two samples positive out of 2,888 tested [4]. In this study, we explore the relative sensitivity of sample pools of varying sizes compared to standard single-specimen SARS-CoV-2 rRT-PCR protocols, the relative efficiency gains yielded by different pool sizes, and the effect of disease prevalence on these gains. We have also developed a standard operating procedure and tested its implementation, both in our own busy diagnostic service and in another laboratory. Finally, we summarize our experience with processing over 29,000 diagnostic samples in pools of two different sizes during two periods of peak diagnostic demand.

## 2. METHODS

### 2.1 Study setting

The Victorian Infectious Diseases Reference Laboratory (VIDRL) is a reference laboratory located in Melbourne, Australia. VIDRL performed all of the SARS-CoV-2 diagnostic testing for the state of Victoria in the early days of the pandemic. In anticipation of increasing demand for testing, a pre-implementation study for sample pooling was performed with defined samples. Pooling of clinical specimens was formally performed during two periods of high demand in March and May 2020.

### 2.2 Pre-implementation study

Pool sizes consisting of two, four, and eight pre-defined samples were used for this study. Each pool contained a single known SARS-CoV-2 positive sample from the reference collection at VIDRL; all other specimens were known negatives. The positive samples varied in viral load, having previously tested at a range of cycle threshold (Ct) values ranging from 19.3 to 35.6 in our standard diagnostic RNA-dependent RNA polymerase (RdRp) gene rRT-PCR assay [5]. Eight pools of each size were designed for use in the study.

### 2.3 Diagnostic testing of clinical samples

A total of 10,312 and 19,388 clinical samples were pooled for testing in March and May 2020, respectively. Samples from lower acuity settings (e.g. outpatient clinics) were selected for pooling, whereas samples from hospital inpatients, healthcare workers, and outbreak investigations – representing more urgent and/or higher prevalence settings – were specifically excluded from pooling. A large variety of swabs was received, including dry swabs to which viral transport medium was added during initial specimen processing.

### 2.4 Sample pooling

An equal amount of fluid from each specimen in a pool was combined into a single tube to give a final volume of 800uL (i.e. for a four-sample pool, 200uL was aliquoted from each sample). The mixture was then vortexed for five seconds prior to nucleic acid extraction. Individual samples were retained for further testing should the pool test positive.

### 2.5 Nucleic acid extraction and complementary DNA (cDNA) synthesis

200μl of pooled sample was extracted for viral RNA with the QIAamp 96 Virus QIAcube HT kit (Qiagen, Germany) on the QIAcube HT System (QIAGEN, Germany) according to manufacturer’s instructions. Purified nucleic acid was then immediately converted to cDNA by reverse transcription with random hexamers using the SensiFAST cDNA Synthesis Kit (Bioline Reagents Ltd., UK) according to manufacturer’s instructions. cDNA was used immediately in the rRT-PCR or stored at -20°C.

### 2.6 SARS-CoV-2 rRT-PCR

3ul of cDNA was added to a commercial real-time PCR master mix (Primer Design PecisionFast qPCR Master Mix, UK) in a 20μl reaction mix containing primers and probe with a final concentration of 0.9μM and 0.2μM for each primer and the probe, respectively.

Primary screening of pooled samples was performed with a SARS-CoV-2 rRT-PCR targeting the RdRp gene [5]. RdRp-positive pools were then ‘deconstructed’, with each individual sample within that positive pool undergoing nucleic acid extraction and testing with rRT-PCRs targeting the E and N genes for confirmation [1]. SARS-CoV-2 was reported as ‘detected’ in an individual sample if either the E or N gene was detected, as the RdRp gene detection in the pool could then be attributed to that sample, thereby fulfilling the Australian Public Health Laboratory Network recommendation of two targets for the detection of SARS-CoV-2 [6].

An in-house positive extraction control, a negative control and a positive control were included with each PCR run. Thermal cycling and rRT-PCR analysis for all assays were performed on the ABI 7500 FAST real-time PCR system (Applied Biosystems, Foster City, CA) with the following thermal cycling profile: 95°C for 2 min, followed by 45 PCR cycles of 95°C for 5 sec and 60°C for 25 sec.

### 2.7 Statistical analysis

Assay sensitivity was compared for each pool size used in the pre-implementation study. The delta Ct value (ΔCt) was defined as the absolute increase in Ct value when the pooled sample was tested compared to when the positive sample was tested individually. A positive ΔCt value (i.e. an increase in Ct value in the pooled sample) therefore represents the loss of PCR sensitivity attributable to sample pooling.

The interaction between the prevalence of SARS-CoV-2 in the test population and the efficiency of the various pool sizes was examined for a disease prevalence ranging from 0.2% to 20%. As prevalence rises, the probability of positive pools requiring deconstruction and further individual testing rises with it. The expected number of tests required per 1,000 individuals for pooled testing was calculated based on the method described by Black et al. [7] using the open source R Shiny app (www.chrisbilder.com/shiny). This method requires calculation of the probability of a pooled sample testing positive, and this probability is derived from assay sensitivity and specificity as well as the prevalence of disease. For the purposes of this study, the sensitivity and specificity of the test was assumed to be fixed at 99%.

## 3. RESULTS

### 3.1 Sensitivity of Pool Sizes

The mean Ct of the positive sample in each pool, the mean Ct of the pooled sample, and the mean ΔCt across the different pool sizes are shown in Table I. All positive samples in each pool were successfully identified by retesting of individual samples as described in the study protocol.

**Table I.** Mean ΔCt for each pool size, representing the loss of PCR sensitivity attributable to pooling

| Pool size (N= 8 pools per pool size) | Mean Ct of positive sample (range) | Mean Ct of pooled sample (range) | Mean ΔCt (range) |
| --- | --- | --- | --- |
| Two samples | 30.4 (23.5 to 34.4) | 31.0 (24.1 to 35.5) | 0.6 (-0.1 to 1.3) |
| Four samples | 30.4 (19.3 to 35.6) | 32.7 (21.7 to 37.2) | 2.3 (0.6 to 3.1) |
| Eight samples | 28.8 (23.0 to 35.0) | 31.8 (26.0 to 38.9) | 3.0 (2.3 to 3.9) |

### 3.2 Pooling Logistics

The approximate time taken for pool assembly was 30 minutes for two-sample pools, 60 minutes for four-sample pools, and 120 minutes for eight-sample pools in a standard 96-well format. Oversight of the pool assembly process was provided by a second staff member to mitigate potential laboratory error. After trialling pools of two, four, or eight samples, larger pools were not trialled, as eight-sample pools were considered the upper limit of what was feasible in our laboratory.

### 3.3 Interaction of SARS-CoV-2 prevalence, pool size and efficiency

Efficiency of pooling, expressed as the expected number of PCR reactions required for the testing of 1,000 individual patients – inclusive of initial pool testing and deconstruction of positive pools – varied with disease prevalence and pool size (Table II, Figure 1). The maximum proportion of reagent savings for two, four, and eight sample pooling is 50%, 75%, and 87.5% respectively, hence there are diminishing marginal gains in savings as pool size increases.

**Table II.** expected number of PCR reactions required for testing of 1,000 samples according to pool size and disease prevalence (inclusive of initial pool testing and deconstruction of positive pools)

| Prevalence of COVID-19 (%) | Two-sample pools | Four-sample pools | Eight-sample pools |
| --- | --- | --- | --- |
| 0.2 | 524 | 278 | 160 |
| 0.5 | 530 | 289 | 183 |
| 1 | 539 | 308 | 220 |
| 2 | 558 | 345 | 290 |
| 5 | 614 | 450 | 471 |
| 10 | 706 | 606 | 701 |
| 15 | 789 | 734 | 851 |
| 20 | 871 | 845 | 954 |

**Figure 1.**
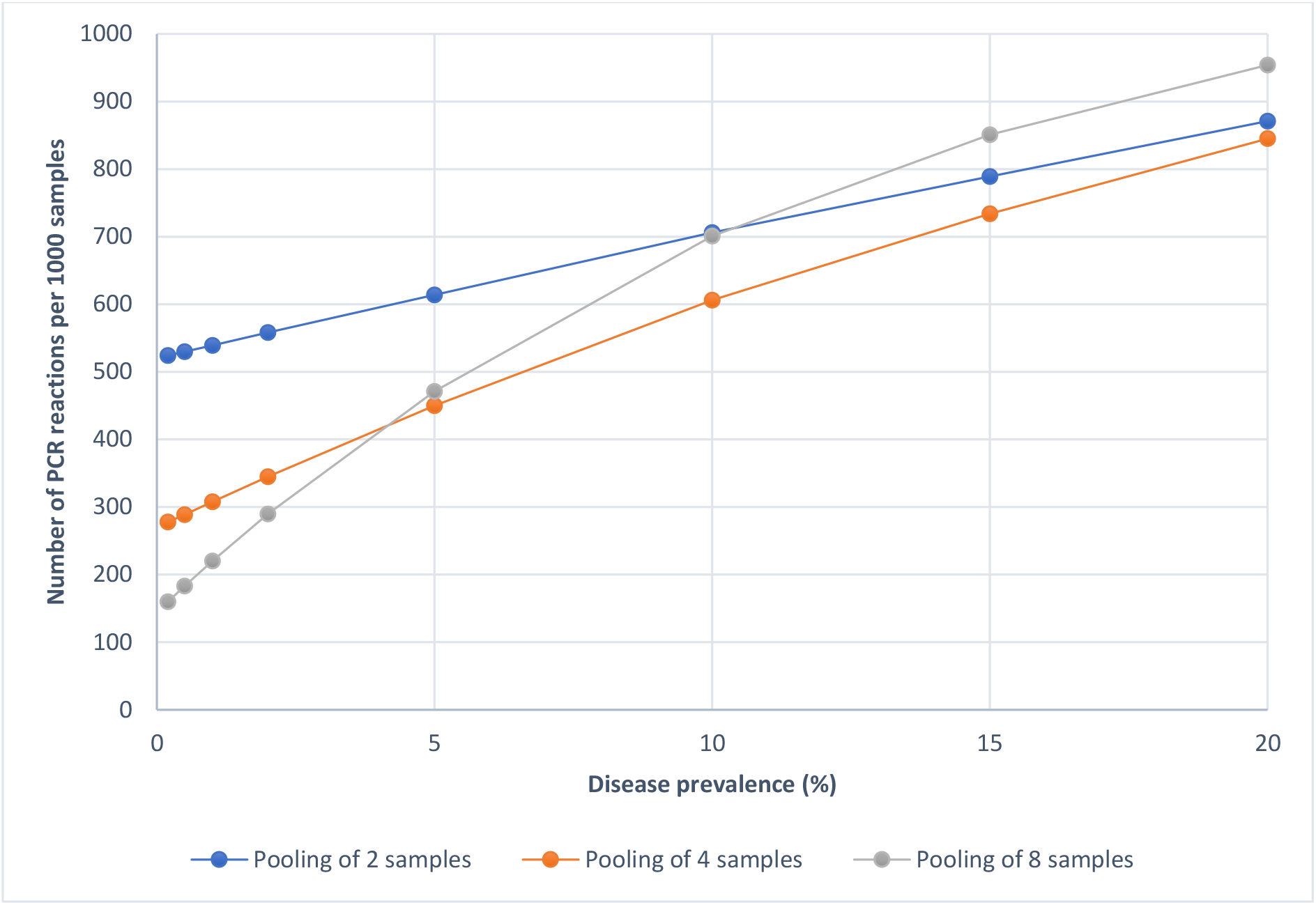
expected number of PCR reactions required per 1,000 samples plotted against disease prevalence

### 3.4 Testing of clinical samples

In March 2020, when the COVID-19 pandemic was in its early stages, and VIDRL was performing diagnostic testing for all of Victoria, testing demand rose to more than 3,000 samples daily. As disease prevalence was <0.5% at the time, pooled testing was performed using eight-sample pools. After a short period of peak demand was managed, our laboratory scaled back to four-sample pools which we considered to offer the best balance of efficiency and sensitivity. The peak daily throughput achieved in this period was 2,222 results reported, compared to a normal daily throughput of approximately 100 respiratory virus RT-PCR tests. Four-sample pooling was again implemented in May 2020, during a high-throughput testing ‘blitz’ targeting low-risk individuals in the community with the goal of detecting asymptomatic cases. A total of 19,388 samples were tested over a 16-day period – 16,609 within seven days, and with a peak day seeing 3,341 results reported. Overall results of these two testing periods is summarised in Table III.

**Table III.** summary of results for pooled testing of clinical samples during two periods of high demand

| Description | Eight-sample pooling<br>(March 2020) | Four-sample pooling<br>(March 2020) | Four-sample pooling<br>(May 2020) |
| --- | --- | --- | --- |
| Pools tested | 250 | 2078 | 4847 |
| RdRp-positive pools | 8 | 49 | 3 |
| Pools with one<br>positive sample | 8 | 41 | 3 |
| Pools with two<br>positive samples | 0 | 8 | 0 |
| Overall positivity rate | 0.4% | 0.6% | 0.02% |

### 3.5 Risk mitigation

Two significant laboratory errors occurred during the first period of high throughput testing using this pooling strategy, and these informed protocol modifications. In each case, error detection and rectification before the next working day obviated adverse clinical impact. Both were human errors; one involving inaccurate manipulation of a sample, and the other being incorrect orientation of a 96-well sample block. The extremely high test throughput and the relative novelty of the pooling protocol were likely cofactors. With pooling, the potential impact of error is magnified by the pooled sample size. Part of our risk mitigation in response to this error was holding back reporting of negative pooled samples on each 96-well block until individual testing and analysis of positive pools was complete.

### 3.6 Inter-laboratory implementation of pooling protocol

Four-sample pooling was successfully implemented by the Microbiological Diagnostic Unit Public Health Laboratory, Doherty Institute (MDU PHL) using the Standard Operating Procedure (SOP) provided by VIDRL (refer to Appendix A for a sample workflow). MDU PHL integrated the pooling process as described, but otherwise retained their in-house testing method – including RNA extraction, PCR process, and gene target – which was different to the method described in this paper. 795 pools representing 3,180 clinical samples were tested during the ‘blitz’ in May 2020. Only one pooled sample tested positive, and the single positive sample in that pool was successfully identified on further testing.

## 4. DISCUSSION

In this study, we demonstrate that pooling is a viable diagnostic strategy for SARS CoV-2 detection in clinical samples, with which relative sparing of nucleic acid extraction kits and PCR reagents could be achieved and test throughput may be significantly increased. Pools of two, four and eight samples could be processed with modified RT-PCR workflows in the context of a very busy diagnostic laboratory setting.

As expected, test sensitivity diminished and testing efficiency increased in proportion to pool size. Two-sample pools were found to impart only a small ΔCt of 0.6, but produced insufficient efficiency gains to justify the workflow change to pooling. Pools of eight produced the greatest efficiency gains at low disease prevalences (0 to 5%) likely to be seen in the early phases of an outbreak, but the significant mean ΔCt of 3.0 suggests that this strategy should be reserved for populations with extremely low pre-test probabilities, such as the asymptomatic general population, and only when testing is required at a scale too high to be achievable by other means. The additional workload of individually testing samples from positive pools of this size becomes impractical in high-prevalence settings. Pools of four appear to provide an efficiency gain over the whole range of prevalences up to 20%, however the gains diminish above 10% prevalence. It would appear that four-sample pools may be considered even when reagents are plentiful, as part of a dedicated workflow for testing large numbers of samples with very low pre-test probability, provided these can be reliably separated from samples requiring tests with maximum sensitivity due to patient disease severity, vulnerability, or the public health risk they potentially represent.

Pool assembly added a complex step to the test workflow, and required focus and precision from staff. To mitigate the potential for laboratory error, we found it was best managed in teams of two experienced staff – one performs the aliquoting of samples, while the other one provides oversight and assembles the individual pools in separate racks. Even so, we experienced human errors during initial implementation, necessitating a change in our reporting protocol as discussed above. Pooling will typically be implemented in the context of high test demand, increasing the risk of error. Consequence of error also potentially increases due to involvement of multiple samples in each pool. Hence risk mitigation is an important part of pooling implementation planning.

Pooling significantly increases the total test throughput achievable in a working day – up to 3,341 in our study. However, the turnaround time for any individual sample is slower via a pooled workflow than with individual testing, due to additional steps of pool assembly and subsequent individual testing of samples in positive pools. High priority samples may be best managed separately from the pooled workflow if this can be achieved. It was also evident that as pooling increases laboratory throughput, pre-analytical stages in specimen reception and data entry are likely to become rate limiting unless resources are available to proportionally upscale these processes.

This study has several weaknesses. Firstly, it is not possible to design a single generic pooling workflow that is applicable to all laboratories. However, the workflow developed in this study has been successfully implemented in at least one other laboratory (MDU PHL) which used a completely different testing method. This demonstrates the transferability of our workflow. Secondly, the clinical impact of the slight reduction in assay sensitivity imparted by pooling was not formally assessed in this study. However, samples with low pre-test probability were specifically selected for pooling, and it is not possible to fully evaluate the false-negativity rate of pooling without large scale parallel testing of individual samples. Additionally, upon reviewing data from the first pooling period in March 2020, we found no instances where a patient with a negative result on pooled testing subsequently tested positive on repeat swab within seven days.

## 5. CONCLUSIONS

In this study, we demonstrate that pooling is a viable strategy for SARS-CoV-2 testing in low-prevalence settings, provide a sample workflow, and report on the successful implementation of this workflow across two laboratories during periods of high testing demand.

## Data Availability

No further data is available online.

## 6. ACKNOWLEDGEMENTS

The authors would like to acknowledge all staff at VIDRL and MDU PHL for their work throughout the SARS-CoV-2 pandemic.

